# Joint association of daytime sleep behaviors and dietary quality with the risk of dementia: A large prospective cohort study

**DOI:** 10.1101/2022.12.17.22283611

**Authors:** Xingqi Cao, Jingyun Zhang, Xin Li, Zhenqing Yang, Kaili Sun, Weiran Chen, Yubo Zhu, Qinglin Xu, Jiahui Xu, Liu He, Xueqin Li, Xiao Tan, Lisan Zhang, Zuyun Liu

## Abstract

**Background:** Identifying modifiable risk factors of dementia is crucial for its early prevention. Daytime sleep behaviors (DSB) and diet are synergetic factors, both potentially important in maintaining cognitive health. However, whether they may jointly influence the risk of dementia in the general population remains unclear.

**Methods:** This study included 187,078 participants from the UK Biobank. DSB burden (low, medium, and high) was assessed through two questions regarding daytime sleepiness and napping. The Recommended Food Score (RFS) was calculated to assess dietary quality (by tertile). We ascertained incident dementia through linkage to hospital inpatient records. We used Cox proportional hazards regression models to estimate the associations.

**Results:** During a median follow-up of 10 years, we documented 1,351 cases with incident dementia. Compared with participants with low DSB burden, those with high DSB burden had a 21% (hazard ratio [HR]: 1.21; 95% confidence interval [CI]: 1.06, 1.38) higher risk of incident dementia. Dietary quality was negatively associated with dementia risk (HR for tertile 1 versus tertile 3: 1.42; 95% CI: 1.24, 1.63). There was a significant interaction between DSB burden and RFS (P for interaction = 0.027). Compared with participants with low DSB burden & high RFS, those with high DSB burden had higher risks of incident dementia, regardless of RFS level.

**Conclusions:** High DSB burden and low dietary quality separately or in joint were significantly associated with higher risks of incident dementia, while high DSB burden seems to play a decisive role. Our findings imply that programs targeting the two behavioral factors, particularly the DSB, may help to prevent dementia.

## Introduction

Dementia, characterized by complex cognitive decline [1], has become one of the major causes of death and dependence, affecting over 55 million people worldwide [2]. Because of the substantial burden not only for the individual with dementia, but also for their caregivers, families, and society [3], dementia has been raised as a public health priority. The absence of curative treatment for dementia [4] has prompted researchers to concentrate on identifying its modifiable risk factors to strengthen its early prevention [5].

Sleep plays a critical role in circadian rhythms, which is closely associated with neurodegenerative diseases [6]. Daytime sleep behaviors (DSB) such as excessive daytime sleepiness and napping are consequences associated with sleep disturbances, which are highly prevalent in older adults [7]. Several cross-sectional and prospective cohort studies have reported the positive associations of daytime sleepiness and napping with dementia [8-15]. Besides, dietary habit is one of the acknowledged modifiable risk factors involved in the development of dementia. Nutritional interventions such as healthy dietary patterns have been recommended by the World Health Organization to prevent or delay cognitive decline and dementia [5]. A recent review suggested that higher adherence to a healthy diet is associated with a lower risk of dementia [16]. However, there is no consensus regarding the efficacy of diet intervention on cognitive function in randomized trials [17-19]. The inconsistent findings may be partially explained by other factors related to dietary quality across the studies. For instance, sleep disturbances are found to alter individuals’ dietary behaviors by disturbing the normal leptin and ghrelin secretions [20, 21], thereby prolonging the time for intake, and resulting in a high intake of energy and fat and a decreased intake of healthy foods (i.e., unhealthy dietary pattern) [22]. On the other hand, some dietary patterns (e.g., low carbohydrate diet, and modest energy restriction diet), foods (e.g., tart cherries, and kiwifruit), and nutrients (e.g., thiamine, folate, and potassium) present promise to improve sleep quality [23-25]. The close interrelationships between sleep and diet imply that DSB and dietary quality may work simultaneously on the development of dementia. Although the separate effects of DSB and dietary quality on dementia have been documented, there is a knowledge gap regarding the joint effect of these two modifiable behavioral factors on the risk of incident dementia.

Using a large sample of the UK Biobank (UKB), we measured dietary quality through the Recommended Food Score (RFS), which is a food-based score as recommended by the dietary guideline [26]. In this study, we aimed to assess the separate and joint effects of DSB burden and dietary quality on the risk of incident dementia over 1,962,646 person-years of follow-up, controlling for a series of potential confounding factors, including genetic susceptibility (i.e., APOE genotypes).

## Methods Participants

This prospective cohort study was based on the UKB, a population-based cohort recruiting more than 500,000 adults (aged 40-69 years) between 2006 and 2010 from 22 assessment centers across England, Scotland, and Wales (http://www.ukbiobank.ac.uk/). Among the participants in the UKB, 210,971 participants completed a 24-hour dietary recall questionnaire at least once were considered. Then, 23,893 participants were excluded because they were diagnosed with dementia at the first time of the 24-hour dietary recall questionnaire survey (N = 75), not British white (N = 8,011), had missing data on DSB burden (N = 623), had implausible energy intake (N = 5,578; men < 800 or > 4200 kcal/day, and women < 500 or > 3500 kcal/day [27]), and had no data on APOE genotypes (N = 4,388), and covariates (N = 5,218). Finally, 187,078 participants were included in the analyses.

### DSB burden

As reported in a previous study [28], two self-reported items (i.e., daytime sleepiness and daytime napping) at the baseline survey were used to define the DSB burden. Daytime sleepiness was assessed via the question “How likely are you to doze off or fall asleep during the daytime when you don’t mean to? (e.g., when working, reading, or driving)” with responses of “never/rarely”, “sometimes”, “often”, and “all the time”. Daytime napping was assessed via the question “Do you have a nap during the day” with responses of “never/rarely”, “sometimes”, and “usually”. It was re-coded as missing when participants answered “do not know” or “prefer not to answer” for both questions. Daytime sleepiness was closely correlated with daytime napping with a spearman correlation coefficient of 0.316 (P < 0.001) in UKB. A participant’s DSB burden was defined as high if he/she answered “sometimes” for both two questions or “often/usually/all the time” for at least one of them, medium if he/she answered “sometimes” for one question and “never/rarely” for the other, or low if he/she answered “never/rarely” for both two questions [28].

### RFS

In the UKB, the data of 24-hour dietary recall of the previous day was measured by the Oxford Web-based questionnaire (https://biobank.ndph.ox.ac.uk/showcase/ukb/docs/DietWebQ.pdf), which was first introduced during the recruitment for the last 70,000 participants. The UKB invited participants with valid e-mail addresses to complete the questionnaire from Feb 2011 to June 2012 on four separate occasions. The RFS was developed by Kant et al to measure overall dietary quality [26]. It was calculated based on data from 21 food components collected through 24-hour dietary recall in this study. Each food component was scored 1 if the consumption was above the minimum threshold (15/g per day for non-beverages or 30/g per day for beverages), or 0 otherwise (**Table S1**). Finally, the RFS ranged from 0 to 21, where a higher score indicates better dietary quality. The dietary quality of a participant was classified into low, medium, and high according to the tertiles of RFS.

### Incident dementia

Incident dementia was ascertained through linkage to hospital inpatient records using the International Classification of Diseases (ICD) codes (ICD-9: 290.2, 290.3, 290.4, 291.2, 294.1, 331.0, 331.1, 331.2, 331.5; ICD-10: A81.0, F00, F01, F02, F03, F05.1, F10.6, G30, G31.0, G31.1, G31.8, I67.3) [29]. The time-to-event for each participant was defined as the period from the date of the first response to the 24-hour recall dietary questionnaire to the date of the first diagnosis of dementia, death, loss to follow-up, or Aug 29, 2021, whichever came first.

### Covariates

Several covariates were considered in our study, including age, sex, educational level, Townsend deprivation index (TDI), body mass index (BMI), smoking status, alcohol consumption, regular physical activity, time spent watching TV, sleep duration, insomnia, family history of Alzheimer’s disease, and APOE genotypes. Educational level was classified as high (i.e., college or university degree), intermediate (i.e., A/AS levels or equivalent, O levels/GCSEs or equivalent), and low (i.e., none of the aforementioned) [30]. TDI represented a participant’s socioeconomic status at the area level and was calculated based on the national census output area according to the postcode of each participant [31]. BMI (kg/m^2^) was calculated as the weight (kg) divided by the square of the height (m), and classified into underweight (< 18.5 kg/m^2^), normal (18.5-24.9 kg/m^2^), overweight (25.0-29.9 kg/m^2^), and obesity (≥ 30.0 kg/m^2^). Smoking status was classified as never, previous, and current smoker. Alcohol consumption was classified as none (i.e., never or special occasions only), low (i.e., one to three times per month), intermediate (one to four times per week), and high (daily or almost daily). A participant was defined as having regular physical activity if he/she met the current global health recommendations for physical activity (i.e., 150 minutes of moderate activity or 75 minutes of vigorous activity or an equivalent combination), which equated to ≥ 500 metabolic equivalent-minutes/week [31]. Time spent watching TV was obtained from a self-reported question “In a typical day, how many hours do you spend watching TV?”, and “less than one hour” was defined as 0 hour. Average sleep duration (hours) per day was self-reported. Insomnia was defined as present if participants reported having trouble falling asleep at night or waking up in the middle of the night. Family history of Alzheimer’s disease was classified into yes, and no. APOE genotypes was defined based on two single nucleotide polymorphisms (i.e., *rs7412* and *rs429358*), and classified into ε4-carrier and ε4-noncarrier according to a previous study [32].

### Statistical analysis

Basic characteristics were described by mean (standard deviation [SD]) and number (percentage), and compared across participants with or without incident dementia by t-test and Chi-square test for continuous and categorical variables, respectively.

To investigate the associations of DSB burden with the risk of incident dementia, Cox proportional hazards regression models were conducted. Hazard ratios (HRs) and 95% confidence intervals (CIs) were documented, and the group of a low DSB burden was considered as the reference. Three models were conducted: model 1 was adjusted for age and sex; model 2 was further adjusted for educational level, TDI, BMI, smoking status, alcohol consumption, regular physical activity, time spent watching TV, sleep duration, insomnia, family history of Alzheimer’s disease, and APOE genotypes based on model 1; model 3 was further adjusted for RFS based on model 2. Also, the associations of RFS with the risk of incident dementia were estimated using the fully adjusted Cox proportional hazard regression model. The high level of RFS was set as the reference.

Next, we investigated the joint associations of DSB burden and RFS with the risk of incident dementia. The associations of RFS with the risk of incident dementia stratified by DSB burden, as well as the interaction between DSB burden and RFS were tested at first. Then, we created a dummy variable based on the joint exposures of DSB burden and RFS: low DSB burden & high RFS, low DSB burden & medium RFS, low DSB burden & low RFS, medium DSB burden & high RFS, medium DSB burden & medium RFS, medium DSB burden & low RFS, high DSB burden & high RFS, high DSB burden & medium RFS, and high DSB burden & low RFS. The fully adjusted Cox proportional hazards regression model was used to estimate the joint effect of DSB burden and RFS on the risk of incident dementia with low DSB burden & high RFS as the reference.

Stratified analyses according to sex and APOE genotypes were conducted. The P values for the product terms between exposures and stratification factors were used to estimate the statistical significance of interactions.

To estimate the robustness of the associations, we further conducted several sensitivity analyses. First, to reduce the potential reverse causation, we excluded participants who were followed up for less than 2 years. Second, to estimate the associations among relatively healthy participants, we excluded participants with diagnosed diabetes mellitus (DM), cardiovascular disease (CVD), cancer, and depression at the first time of the dietary quality assessment. Third, to avoid the potential bias due to social isolation, we set the end of follow-up time before the COVID-19 pandemic (i.e., December 2019). Fourth, to account for the competing risk from all-cause death, we used the Fine-Gray model [33]. Fifth, to account for the influence of cardiometabolic-related biomarkers, we further adjusted for glycated hemoglobin A1c (HbA1c), high-density lipoprotein cholesterol (HDL-C), systolic blood pressure (SBP), and diastolic blood pressure (DBP). Sixth, to account for the influence of sleep apnea, we further adjusted for self-reported sleep apnea (yes or no).

All statistical analyses were performed using SAS (SAS Institute, Cary, NC, USA) and R (version 4.1.2), and statistical significance was defined as a two-tailed P < 0.05.

## Results

### Participants characteristics

Of 187,078 participants, 1,351 developed dementia during the 10-year follow-up. In comparison to participants without incident dementia, those who developed dementia were older, and more likely to be male, obese, and less educated. In addition, they tended to have a low level of RFS, and a high level of DSB burden. More details of basic characteristics are shown in **Table 1**.

**Table 1:**
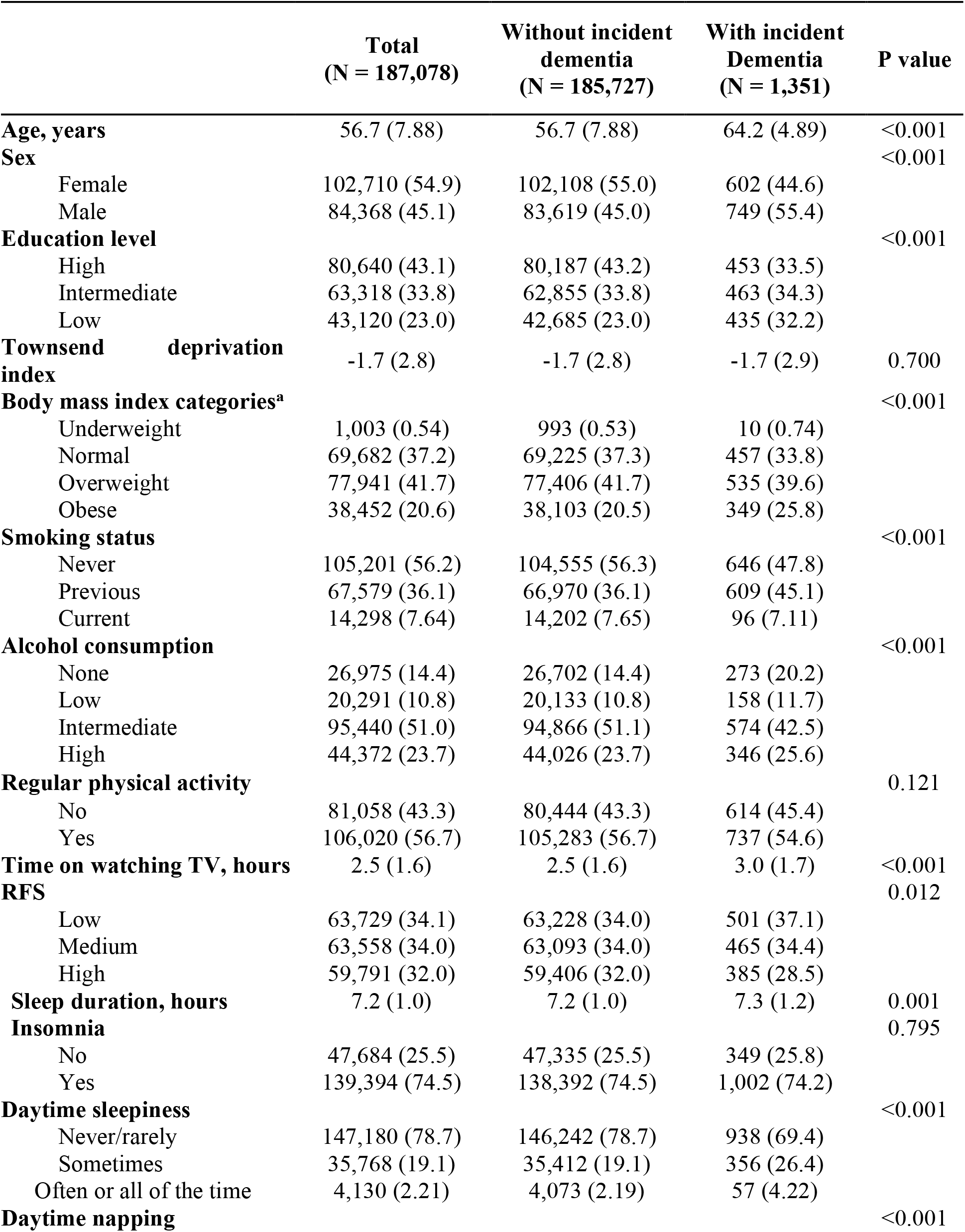
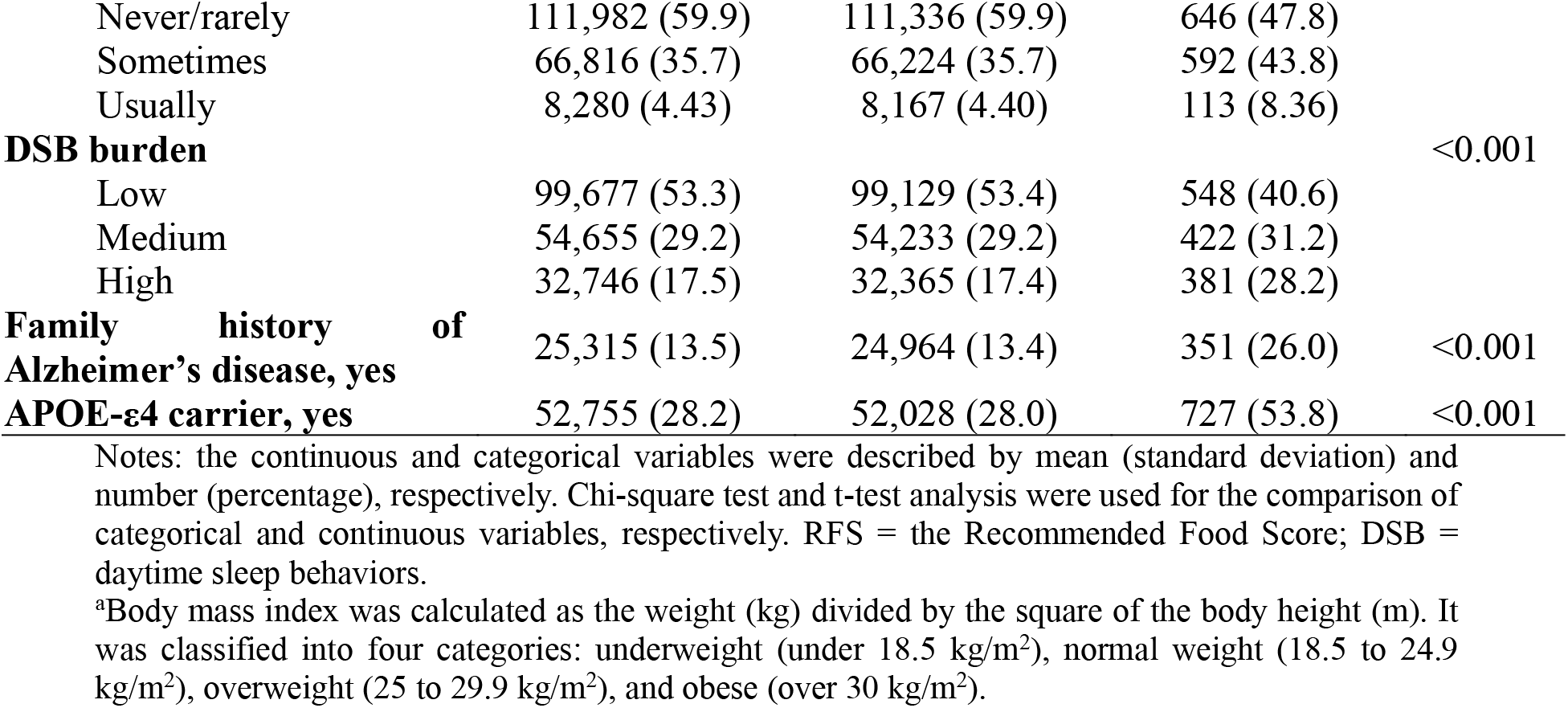
Baseline characteristics of 187,078 participants from UK Biobank.

### Separate associations of DSB burden and RFS with the risk of incident dementia

We observed that DSB burden was positively associated with a higher risk of incident dementia (P for trend < 0.05) (**Table 2**). After adjusting for age, sex, educational level, TDI, BMI, smoking status, alcohol consumption, regular physical activity, time spent watching TV, sleep duration, insomnia, family history of Alzheimer’s disease, and APOE genotypes, when participants with high DSB burden were compared with those with low DSB burden, the risk of incident dementia increased by 21% (HR: 1.21; 95% CI: 1.06, 1.39). After additionally adjusting for RFS, the significant associations remained (HR: 1.21; 95% CI: 1.06, 1.38). Besides, lower RFS was significantly associated with higher risks of incident dementia (**Figure 1**). Compared with participants with high RFS, those with medium, and low RFS had increased risks of incident dementia, with HRs of 1.22 (95% CI: 1.07, 1.40), and 1.42 (1.24, 1.63), respectively (**Table S2**).

**Table 2.** Association between daytime sleep behaviors burden and incident dementia.

|  | HR (95% CI) |  |  | P for trend |
| --- | --- | --- | --- | --- |
|  | Low DSB burden | Medium DSB burden | High DSB burden |  |
| **Events/Person-years** | 548/1,048,154 | 422/576,259 | 381/341,833 | — |
| **Model 1** | Ref. | 1.10 (0.96, 1.25) | 1.28 (1.12, 1.46) | <0.001 |
| **Model 2** | Ref. | 1.07 (0.94, 1.22) | 1.21 (1.06, 1.39) | 0.006 |
| **Model 3** | Ref. | 1.07 (0.94, 1.22) | 1.21 (1.06, 1.38) | 0.007 |

Notes: Model 1 was adjusted for age and sex; Model 2 was further adjusted for education level, Townsend deprivation index, body mass index, smoking status, alcohol consumption, regular physical activity, time spent watching TV, sleep duration, insomnia, family history of Alzheimer's disease, and APOE genotypes; Model 3 was further adjusted for the Recommend Food Score based on model 2. HR = hazard ratio; CI = confidence interval; DSB = daytime sleep behaviors.

**Figure 1.**
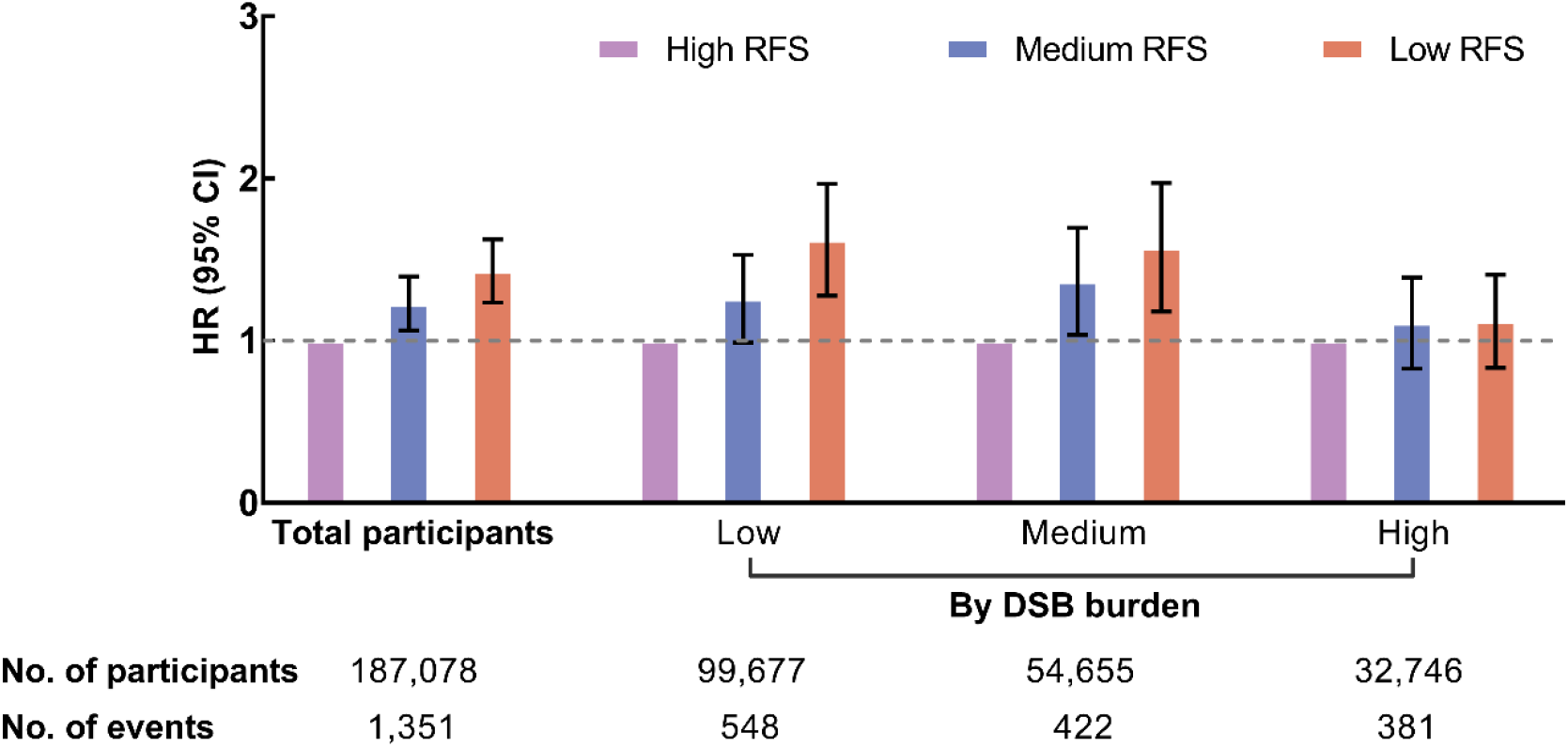
Associations of the Recommend Food Score with incident dementia in total participants and by daytime sleep behaviors burden. The model was adjusted for age, sex, education level, Townsend deprivation index, body mass index, smoking status, alcohol consumption, regular physical activity, time spent watching TV, sleep duration, insomnia, family history of Alzheimer’s disease, APOE genotypes, and DSB burden (in total participants only). The interactive P value of DSB burden and RFS was 0.027. DSB = daytime sleep behaviors; RFS = the Recommended Food Score.

### Joint associations of DSB burden and RFS with the risk of incident dementia

A significant interaction was found between DSB burden and RFS on the risk of incident dementia (P for interaction = 0.027, **Figure 1**). The associations of RFS with dementia risk were more prominent in those with low and medium DSB burden. Compared with participants with high RFS, those with low RFS had significantly higher risk of incident dementia in subgroups of low (HR: 1.60; 95% CI: 1.29, 1.98) and medium (HR: 1.54; 95% CI: 1.20, 1.99) DSB burden. While null associations of RFS with dementia risk were observed among those with high DSB burden (**Table S2**).

**Figure 2** presents the joint associations of DSB burden and RFS with dementia risk (P for trend < 0.001). Compared with participants with low DSB burden & high RFS, higher risks of incident dementia were observed in those with low DSB burden & low RFS (HR: 1.61; 95% CI: 1.30, 1.98), medium DSB burden & medium RFS (HR: 1.41; 95% CI: 1.13, 1.77), medium DSB burden & low RFS (HR: 1.61; 95% CI: 1.29, 2.02), high DSB burden & high RFS (HR: 1.44; 95% CI: 1.13, 1.84), high DSB burden & medium RFS (HR: 1.58; 95% CI: 1.25, 1.99), and high DSB burden & low RFS (HR: 1.60; 95% CI: 1.26, 2.03) (**Table S3**).

**Figure 2.**
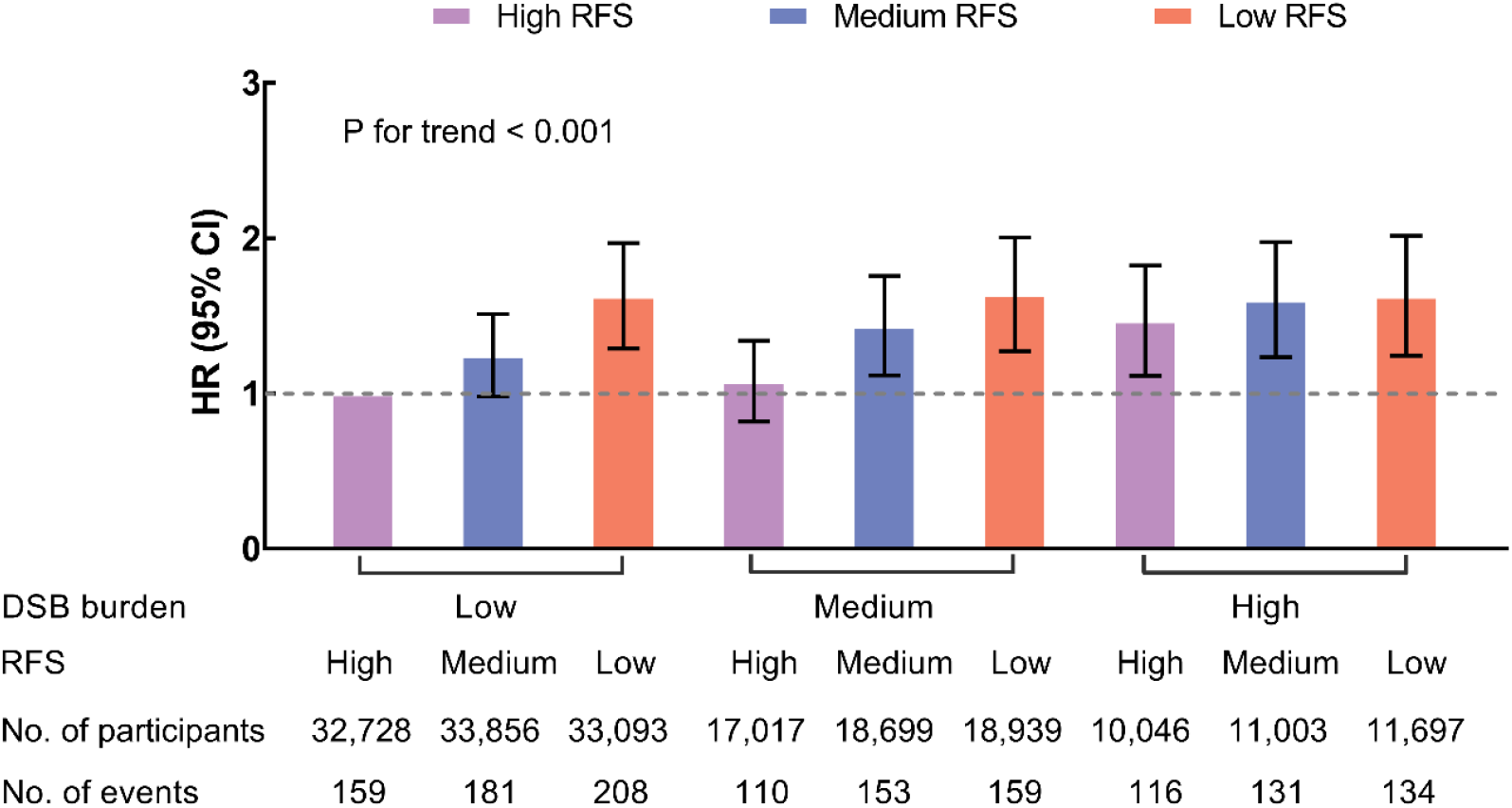
Joint associations of daytime sleep behaviors burden and the Recommend Food Score with incident dementia. The model was adjusted for age, sex, education level, Townsend deprivation index, body mass index, smoking status, alcohol consumption, regular physical activity, time spent watching TV, sleep duration, insomnia, family history of Alzheimer’s disease, and APOE genotypes. HR = hazard ratio; CI = confidence interval; DSB = daytime sleep behaviors; RFS = the Recommended Food Score.

### Stratification and sensitivity analyses

When stratifying by sex and APOE genotypes, similar associations of DSB burden (**Table S4**) and RFS (**Table S5**) with the risk of incident dementia were observed. No significant interactions between DSB burden and sex (P for interaction = 0.470), APOE genotypes (P for interaction = 0.967) were found. In addition, we observed consistent joint associations of DSB burden and RFS with dementia risk in subgroups, as well as non-significant interactions (**Table S6**).

In sensitivity analyses (**Table S7-S9**), generally robust results were observed when excluding participants who were followed up for less than 2 years, excluding participants with diagnosed DM, CVD, cancer, and depression at baseline, setting the end of follow-up time before the COVID-19 pandemic (i.e., December 2019), or adjusting for all-cause death as competing risk. In addition, the results did not materially change when further adjusting for cardiometabolic-related biomarkers (i.e., HbA1c, HDL-C, SBP, and DBP), or sleep apnea.

## Discussion

In this large population-based cohort study of adults with 1,962,646 person-years of follow-up, we have three main findings. First, higher DSB burden, as well as lower adherence to RFS, were significantly associated with a higher risk of incident dementia. Second, the associations of RFS with dementia risk were more prominent among those with low and medium levels of DSB burden. Third, there were significant statistical interactions between DSB burden and RFS on dementia risk; the high risk was persistently observed in participants with high DSB burden regardless of RFS. Of note, these associations were independent of APOE genotypes, a known genetic factor of dementia. Our findings highlight the importance of reducing the DSB burden and adhering to a healthy diet, particularly the former, in the prevention of dementia.

Several prospective studies have suggested positive associations of the separate burdens of daytime sleepiness and daytime napping with dementia risk [8-12] with one exception [34]. Thus, our finding that a higher DSB burden increases the risk of incident dementia is consistent with these results. Furthermore, nighttime sleep disturbances are associated with higher dementia risk [35-38]. A high DSB burden may reflect poor sleep quality at night. We adjusted for common nighttime sleep characteristics (i.e., sleep duration and insomnia), and found that our results remained to be significant. Also, self-reported sleep apnea did not influence the observed associations. These findings imply an independent role of DSB burden from nighttime sleep patterns. Research is warranted to clarify the separate underlying mechanisms of DSB burden and night sleep disturbances in the development of dementia. On the other hand, daytime sleepiness and napping are found to be linked with brain β-amyloid deposition in adults without dementia [39, 40]. Together with findings of the bidirectional associations of daytime napping with Alzheimer’s dementia from a recent study [12], we speculate that increased DSB burden might be the early symptom that emerged at the preclinical stage of dementia. Hence, early assessments and interventions of DSB burden may help identify vulnerable individuals, delay the progression of dementia, and further reduce the dementia-related burden.

Our findings of the negative associations between dietary quality and dementia risk are consistent with results from previous studies concerning the Mediterranean diet [41], Dietary Approaches to Stop Hypertension [42], and Mediterranean-DASH Intervention for Neurodegenerative Delay diet [42-44]. In this study, we used RFS, a food-based score that focused more on healthy food consumption and was relatively less affected by reporting bias of portion size. Intriguingly, we further observed that another behavioral factor, DSB burden, modified the associations of dietary quality with dementia risk, underscoring the importance of DSB burden in determining the associations between dietary quality and subsequent dementia. Poor sleep quality is often associated with more frequent nighttime awakening, which further results in a high DSB burden. Evidence suggests that inadequate sleep may lead to poor dietary quality by increased ghrelin/decreased leptin [21], followed by an increased hedonic drive for foods [45] and altered taste sensitivity or perception [45]. Conversely, dietary quality may regulate hormones and inflammation status that are linked to sleep quality [46]. As a consequence, the DSB burden may interact with dietary quality, contributing to the increased risk of dementia. Although these observations are biologically plausible, further studies are required to examine the underlying mechanisms of DSB burden and dietary quality in developing dementia.

The associations of the combination of DSB burden and RFS on the risk of incident dementia were supported by a recent study from the China Health and Nutrition Survey [47]. Shang et al. conducted a longitudinal study in 2,307 middle-aged and older adults, and found that adults with both healthy sleep and dietary pattern had a 37% lower risk of cognitive decline [47]. To our knowledge, the present study provided the first evidence that joint exposure to either high DSB burden or low RFS level increased the risk of incident dementia. We found that individuals with a low DSB burden could lose their protection if they had poor dietary quality. Although experimental studies are required to verify our findings, there may be the potential to delay the progression of dementia by regulating dietary habits when experiencing relatively lower DSB burden. On the other hand, the current study observed that participants with high DSB burden persistently show a high risk of dementia regardless of the level of RFS. As suggested, improving dietary quality may be futile if the DSB burden could not be reduced, preventive strategies targeting the DSB burden should be prioritized over dietary interventions. Furthermore, considering that DSB may reflect the preclinical symptom of dementia [12], the assessment of DSB burden at the early stage may help identify vulnerable individuals. Altogether, the results support that the integrated management of behaviors (i.e., DSB and diet quality), with DSB as the predominant, might be an effective strategy to prevent or delay the development of dementia.

## Strength and limitation

The main strengths of this study include the relatively large sample size, as well as the design of a prospective cohort study with a median follow-up of 10 years. Also, several limitations of this study should be pointed out. First, single assessments of DSB burden and RFS were obtained, and there was a time gap between DSB and RFS assessments; however, behaviors might change over time, which may influence our risk estimation. Future research on the effects of long-term changes in DSB burden and dietary quality on dementia risk is required. Second, the DSB burden was measured subjectively. Although polysomnography is the gold standard for sleep assessment, it is not feasible in large-scale population-based studies. Other objective assessment techniques, such as actigraphy, will allow for a better understanding of the complexity of DSB. Third, the dietary information was self-reported; so recall bias was inevitable, and RFS may not fully reflect dietary intake. Fourth, UKB participants were relatively healthier compared to the general population in the UK, and our sample only included White, which limited the generalizability of the findings. More studies in other populations are warranted to verify our findings. Finally, due to the observational study design, the causal associations between DSB burden, diet, and dementia risk cannot be determined. Future studies are needed to estimate whether interventions targeting at DSB and diet can reduce the risk of dementia.

## Conclusions

In summary, based on a large population-based cohort study from the UK, this study provided evidence that high DSB burden and low adherence to RFS separately increased the risk of incident dementia. Furthermore, high DSB burden was persistently associated with a high risk of incident dementia, regardless of RFS level. Our findings underscore the importance of monitoring the DSB burden and dietary quality, particularly the former, for the early detection and prevention of dementia. Integrated management of behaviors, including improving DSB and dietary quality, may contribute to the alleviation of the dementia burden.

## Data Availability

The datasets supporting the conclusions of this article are available at www.ukbiobank.ac.uk/register-apply.

https://www.ukbiobank.ac.uk/enable-your-research/register

## Author contributions

LZ and ZL designed and coordinated the study. XC and JZ analyzed the data and drafted the article. Xin Li, ZY, KS, WC, YZ, QX, JX, LH, Xueqin Li, XT, LZ, and ZL contributed to the interpretation of data. Xin Li, ZY, KS, WC, YZ, QX, JX, LH, Xueqin Li, XT, LZ, and ZL revised the manuscript. All authors read and approved the final version of the manuscript.

## Acknowledgments

This research has been conducted using the UKB resource under application number 61856. We wish to acknowledge the UK Biobank participants who provided the sample that made the data available.

